# Telehealth in Rheumatology: The 2021 Arab League of Rheumatology Best Practice Guidelines

**DOI:** 10.1101/2021.08.18.21262218

**Authors:** Nelly Ziade, Ihsane Hmamouchi, Lina el Kibbi, Melissa Daou, Nizar Abdulateef, Fatemah Abutiban, Bassel Elzorkany, Chafia Dahou-Makhloufi, Wafa Hamdi, Samar Al Emadi, Hussein Halabi, Khalid A. Alnaqbi, Sima Abu Al Saoud, Soad Hashad, Radouane Niamane, Manal El Rakawi, Layla Kazkaz, Sahar Saad, Mervat Eissa, Ilanca Fraser, Basel Masri

## Abstract

**Background:** Telehealth use is increasing and will undeniably continue to play a role beyond the COVID-19 era. Best practice guidelines (BPG) for telehealth add credibility, standardize approaches, facilitate reimbursement, and decrease liability.

**Objectives:** To develop BPG for the use of Telehealth In Rheumatology in the Arab region, to identify the top barriers and facilitators of telehealth in the Arab region, and to provide rheumatologists with a practical toolkit for the implementation of telehealth.

**Methods:** Guidelines were drafted by a core steering committee from the Arab League of Associations for Rheumatology (ArLAR) after performing a literature search. A multidisciplinary task force (TF), including 18 rheumatologists, 2 patients, and 2 regulators from 15 Arab countries, assessed the BPG using 3 rounds of anonymous online voting by modified Delphi process. The voting on barriers and facilitators was performed through one voting round. The toolkit was developed based on available literature and discussions during the Delphi rounds.

**Results:** Four General Principles and twelve Statements were formulated. All statements reached >80% consensus. A teleconsultation was specifically defined for the purpose of these guidelines. The concept of choice in telehealth was highlighted, emphasizing patient confidentiality, medical information security, rheumatologist’s clinical judgment, and local jurisdictional regulations. The top barrier for telehealth was the concern about the quality of care. The toolkit emphasized technical aspects of teleconsultation and proposed a triage system.

**Conclusions:** The ArLAR BPG provides rheumatologists with a series of strategies about the most reliable, productive, and rational approaches to apply telehealth.

**Article Summary:** *Strengths and limitations of this study:*

- Best practice guidelines (BPG) the use of Telehealth In Rheumatology in the Arab region were developed herein under the umbrella of the Arab League of Associations for Rheumatology (ArLAR)
- A teleconsultation was specifically defined for the purpose of these guidelines
- The concept of choice in telehealth was highlighted, emphasizing patient confidentiality, medical information security, rheumatologist’s clinical judgment, and local jurisdictional regulations
- The top barrier for telehealth was the concern about the quality of care
- The ArLAR BPG provides rheumatologists with a series of strategies about the most reliable, productive, and rational approaches to apply telehealth in the rheumatology clinic

## INTRODUCTION

Telehealth services were projected to light with the emergence of the Coronavirus Disease 19 (COVID-19) pandemic in 2020, as the demand for telehealth largely exceeded the supply.[1– 3] Even though this high demand may decline after the pandemic’s end, the need for telehealth services would undeniably persist in the future.

The World Health Organization defines telehealth as *“the offering of health care services, where distance is an essential aspect, by all healthcare practitioners using information and communication technology such as mobile consultation, video calling, written report or protected email, to share valid information for the diagnosis, treatment, and prevention of disease, research and evaluation, and for the continuing education of health care providers, all in the interest of advancing the health of individuals and their communities”*.[4]

The clinical applications of telehealth are wildly diverse, and it is currently available in various settings, including emergency departments, inpatient hospital wards, intensive care units, and commercial pharmacies.[5] In addition, significant advancements in technology have established telehealth as a feasible option for managing patients with rheumatic diseases. Studies indicate how the use of telerheumatology in certain settings has successfully increased access to specialty care, with good patient and provider satisfaction.[6,7]

Surveys conducted by the Arab League of Associations for Rheumatology (ArLAR) to study the impact of the COVID-19 pandemic on patients with chronic rheumatic diseases and rheumatologists in 15 Arab countries identified telehealth as a significant unmet need in the management of these patients.[3,8] The successful implementation of telerheumatology can assist rheumatologists in providing continuity of care for patients who face obstacles that restrict their access to in-person treatment. However, the lack of a structural framework for telerheumatology has been identified as a barrier to the effective implementation of telehealth in the rheumatology clinic [10].[9] Best practice guidelines (BPG) for telerheumatology can provide the necessary framework to facilitate rheumatology teleconsultation as they add credibility, standardize approaches, decrease liability and facilitate reimbursement for this novel health service.[10] Nevertheless, such BPG for performing telehealth services in rheumatology and practical advice for their implementation are still lacking.

The primary objective of this study was to develop BPG for the use of Telehealth In RheumatOLogy (TIROL study) in the Arab region. The secondary objectives were to identify the top barriers and facilitators of telehealth in the ArLAR countries, and to provide rheumatologists with a practical toolkit for the implementation of telehealth in the rheumatology clinic.

## METHODS

The BPG were developed under the umbrella of the ArLAR, in line with the Appraisal of Guidelines for REsearch & Evaluation (AGREE II) instrument.[11] A core steering committee (SC) performed a computerized literature search of four sources (PubMED, American College of Rheumatology (ACR), American Telemedicine Association, and World Health Organization) to identify available guidelines and studies using keywords “telerheumatology” and “telemedicine-guidelines”. The literature search identified 1494 potentially eligible studies. Of these, 50 were ultimately included to formulate the guidelines, identify the barriers and facilitators for telehealth and design the implementation toolkit. Studies varied significantly in publication type, quality of evidence, and methods’ reporting (**Figure 1**).

**FIGURE 1.**
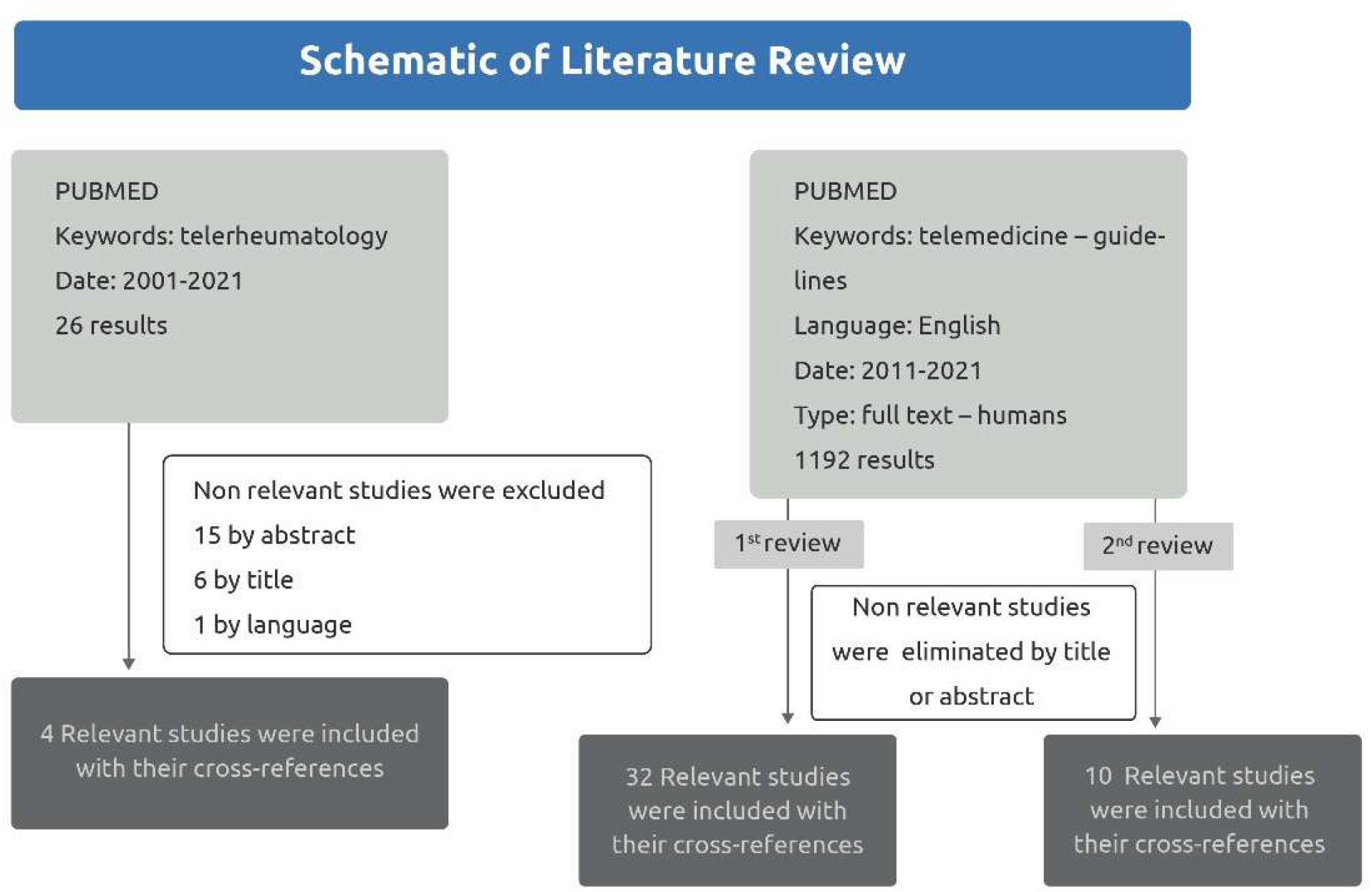
SCHEMATIC OF LITERATURE REVIEW.

Four General Principles (GP) and twelve Best Practice Statements (BPS) were formulated, and levels of evidence were indicated according to the Oxford Centre for Evidence-Based Medicine.[12] The draft was reviewed by the ArLAR scientific committee, a law firm advisor, and an ACR telemedicine expert. Thereafter, a multidisciplinary task force (TF), including rheumatologists (n = 18) (from the Arab Adult Arthritis Awareness (AAAA) group, a special interest group from ArLAR and two ArLAR advisors), patients with rheumatic diseases (n=2), regulators and payers (n=2, one from the public sector and the other from the private sector) from 15 Arab countries convened online (through an online platform) and assessed the BPG using three rounds of voting by modified Delphi process (www.calibrum.com) (**Figure 2**).

**FIGURE 2.**
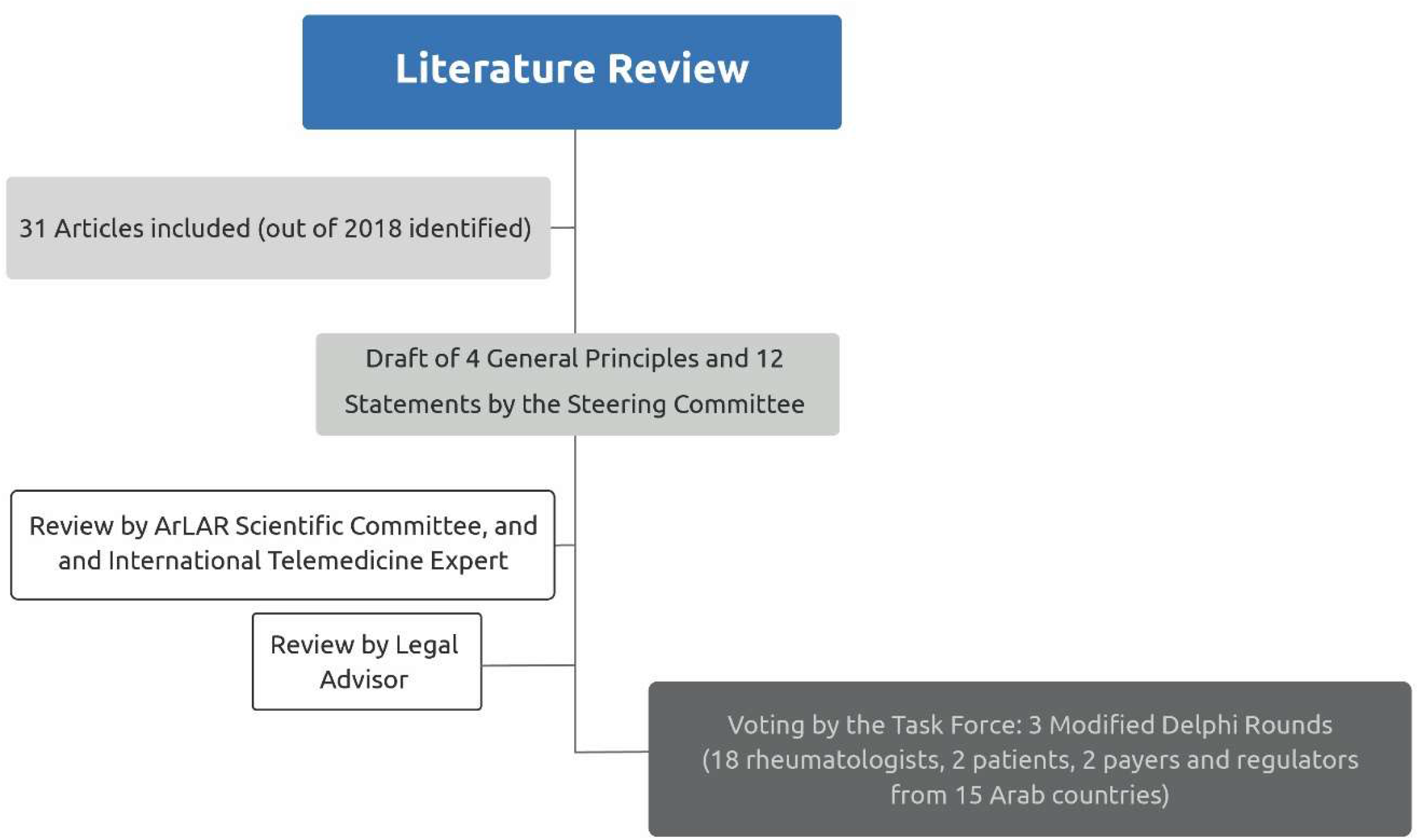
STUDY FLOWCHART.

The levels of evidence and the decision rules were explained to the TF during a briefing meeting two weeks before the first voting round. Taskforce members reported their level of agreement during each round of voting using a numerical rating scale of one (complete disagreement) to nine (complete agreement). Participants were also able to give qualitative comments. The first two Delphi rounds were performed through live online meetings moderated by one member of the steering committee and the last round used asynchronous voting through the Surveylet platform. During each round, results were discussed by all members of the TF. Revisions to the GP and BPS were made by the SC through an iterative process until consensus was achieved. The criteria for moving from one Delphi round to another and for selecting the final statement were guided by the OMERACT recommendations:[13] GP and BPS were included in the final BPG without further voting if ≥ 80% of TF members indicated high agreement (7-9). All GP and BPS that scored ≥ 50% high agreement (7-9) and ≤ 15% low agreement (1-3), all items that required reformulation were included in the next round of voting. The criteria to select a GP and BPS in the final round were ≥ 70% high agreement (7-9) and ≤ 15% low agreement (1-3). All TF members (n=22) participated in each of the 3 rounds. All votes were anonymous and weighted equally.

Possible physician and patient-related barriers and facilitators to telehealth were identified through the literature search. Before evaluating the BPG, TF members were asked to rank the barriers and facilitators of telehealth in rheumatology in the Arab region from one (most important barrier or facilitator, respectively) to five (least important barrier or facilitator, respectively). The voting on barriers and facilitators was done by one round of anonymous voting through the Surveylet platform. Finally, based on the literature review and discussions with the TF during the voting rounds, the SC proposed a practical toolkit for the possible implementation of telehealth in the rheumatology clinic.

### Patient and Public Involvement

Two patients with rheumatic diseases were invited to participate in the Delphi panel.

## RESULTS

The final BPG are presented in **Table 1**, with the accompanying level of evidence (LoE), consensus, and level of agreement (LoA) for each round of voting. All GP and BPS reached >80% consensus by the end of round 3 **(Supplementary Table 1)**.

**Table 1.**
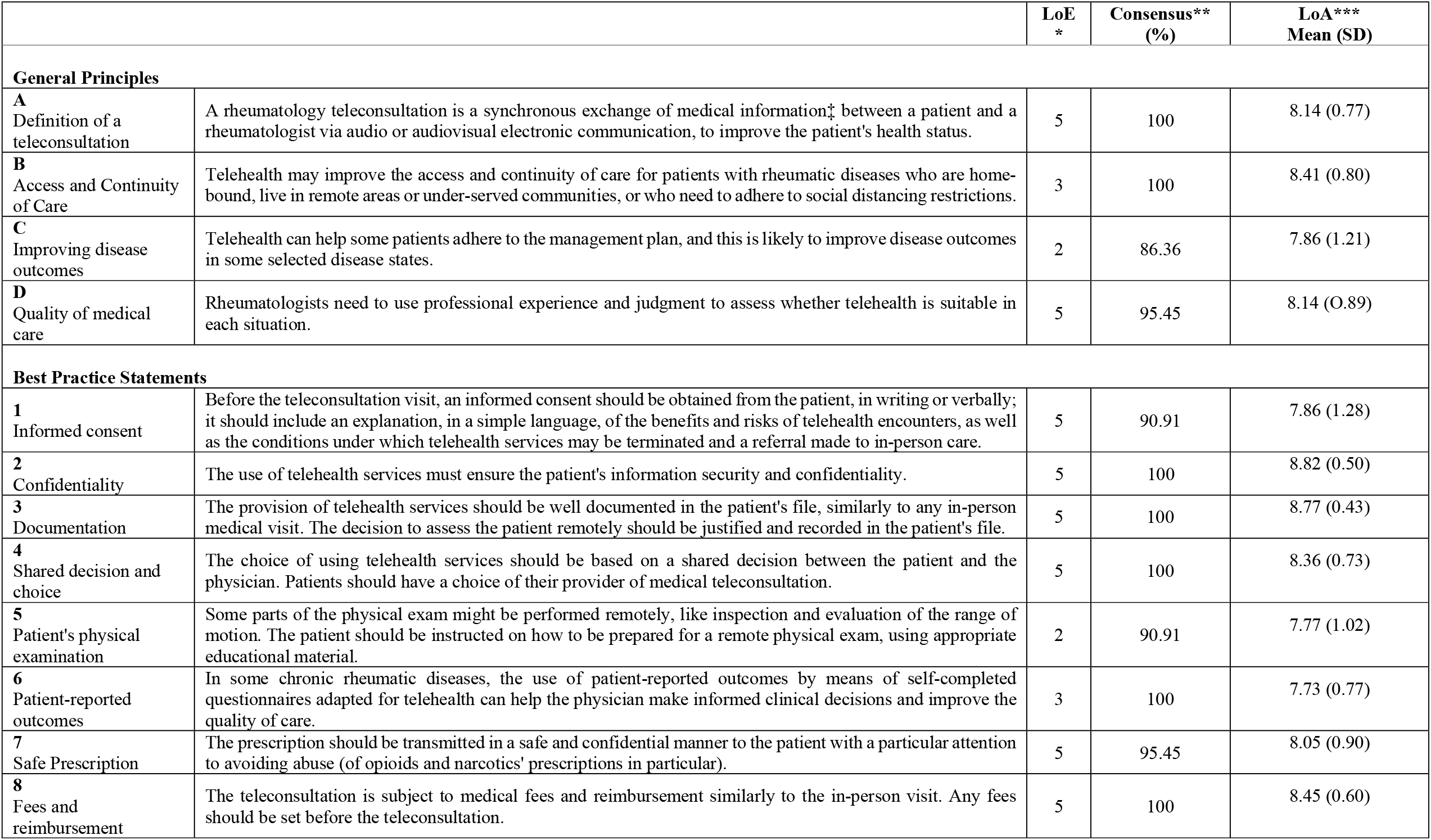
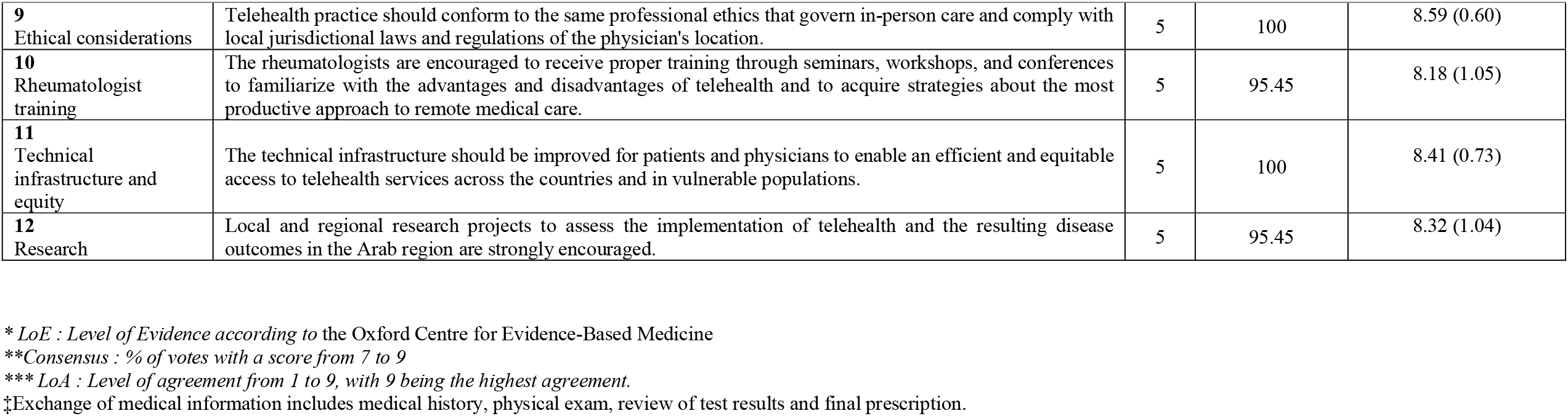
General Principles and Best Practice Guidelines Statements for Telehealth in Rheumatology.

### General Principles

#### General principle A: Definition of a teleconsultation (LoE 5; LoA 8.14; Consensus 100%)

Adapted from the American Telemedicine Association’s definition of “telemedicine” [5,13,14] the definition of a teleconsultation was established specifically for these guidelines as: “*A rheumatology teleconsultation is a synchronous exchange of medical information between a patient and a rheumatologist via audio or audiovisual electronic communication, to improve the patient’s health status”*, where “medical information” includes medical history, physical exam, review of test results and the final prescription and “synchronous” refers to a real-time exchange between the patient and the physician via video, audio or text. [15]

The TF is aware that the definition of teleconsultation will likely vary in each country. For example, under United Arab Emirates laws, audio communication by phone or mobile would be part of the electronic communication used for teleconsultation.[16]

Notably, teleconsultation is differentiated from e-consultation, which is an exchange of medical information between two healthcare providers via electronic audiovisual communication to improve a patient’s health status. Therefore, E-consultations are not within the scope of the current BPG.

#### General principle B: Access and Continuity of Care (LoE 3; LoA 8.41; Consensus 100%)

*Telehealth may improve the access and continuity of care for patients with rheumatic diseases who are home-bound, live in remote areas or under-served communities, or who need to adhere to social distancing restrictions*.

This principle is supported by the ACR Position statement on telemedicine[17] and is especially true in the era of COVID-19. Studies have shown that measures related to the containment of the COVID-19 pandemic, like national lockdowns and social distancing restrictions, led to a perceived delay between symptom onset and a first rheumatological visit.[2] Several international surveys also found that, during the COVID-19 pandemic, 10% to 25% of patients with rheumatic diseases stopped their chronic medication, thus compromising the treat-to-target strategies and emphasizing the need for an alternative to in-clinic visits.[1–3,18]

This principle is also supported by the results from a survey conducted with 75 rheumatologists in the Netherlands during the COVID-19 pandemic, which found that continuity of care was guaranteed through telephone and video consultations by 99% and 9% of the respondents, respectively.[19] Another survey conducted with 426 established patients and 74 physicians found that virtual video visits (VVVs) were vastly preferred to office visits by patients for convenience and travel time, while the majority (52.5%) of clinicians reported higher efficiency of a VVV appointment.[20]

Evidence from recent research, therefore, suggests that when implemented correctly, telehealth may provide a well-accepted way of remote consultation with patients with rheumatic diseases and provide good patient satisfaction regarding access and continuity of care.[6,19,21] It is, however, imperative that periodic in-person visits continue to form part of patients’ ongoing care.[14]

#### General principle C: Improving disease outcomes (LoE 2; LoA 7.86; Consensus 86.36%)

*Telehealth can help some patients adhere to the management plan, and this is likely to improve disease outcomes in some selected disease states*.

Using a proper triage system for telehealth in rheumatology, early detection, and early referral may improve disease outcomes, especially in diseases like rheumatoid arthritis (RA) where there is a limited window of opportunity for early management. By facilitating the application of the treat-to-target strategy in RA and other chronic diseases (such as gout for example), telehealth is likely to support adherence to therapy and maintaining treatment targets [23-24].[7,22] However, higher-quality randomized controlled trials are needed to demonstrate the effectiveness of different telerheumatology interventions in improving disease outcomes.[21]

#### General principle D: Quality of medical care (LoE 5; LoA 8.14; Consensus 95.45%)

*Rheumatologists need to use professional experience and judgment to assess whether telehealth is suitable in each situation*.

Rheumatologists should use their judgment to decide whether a teleconsultation is appropriate in each case. They should ensure that the quality of care provided remotely via telehealth services is consistent with related in-person services while ensuring that the provided services are in alignment with the local laws and regulations where the rheumatologist is based. A guide for a triage system is proposed in the Toolkit section of this document **(Figure 3)**.[7,22]

**FIGURE 3.**
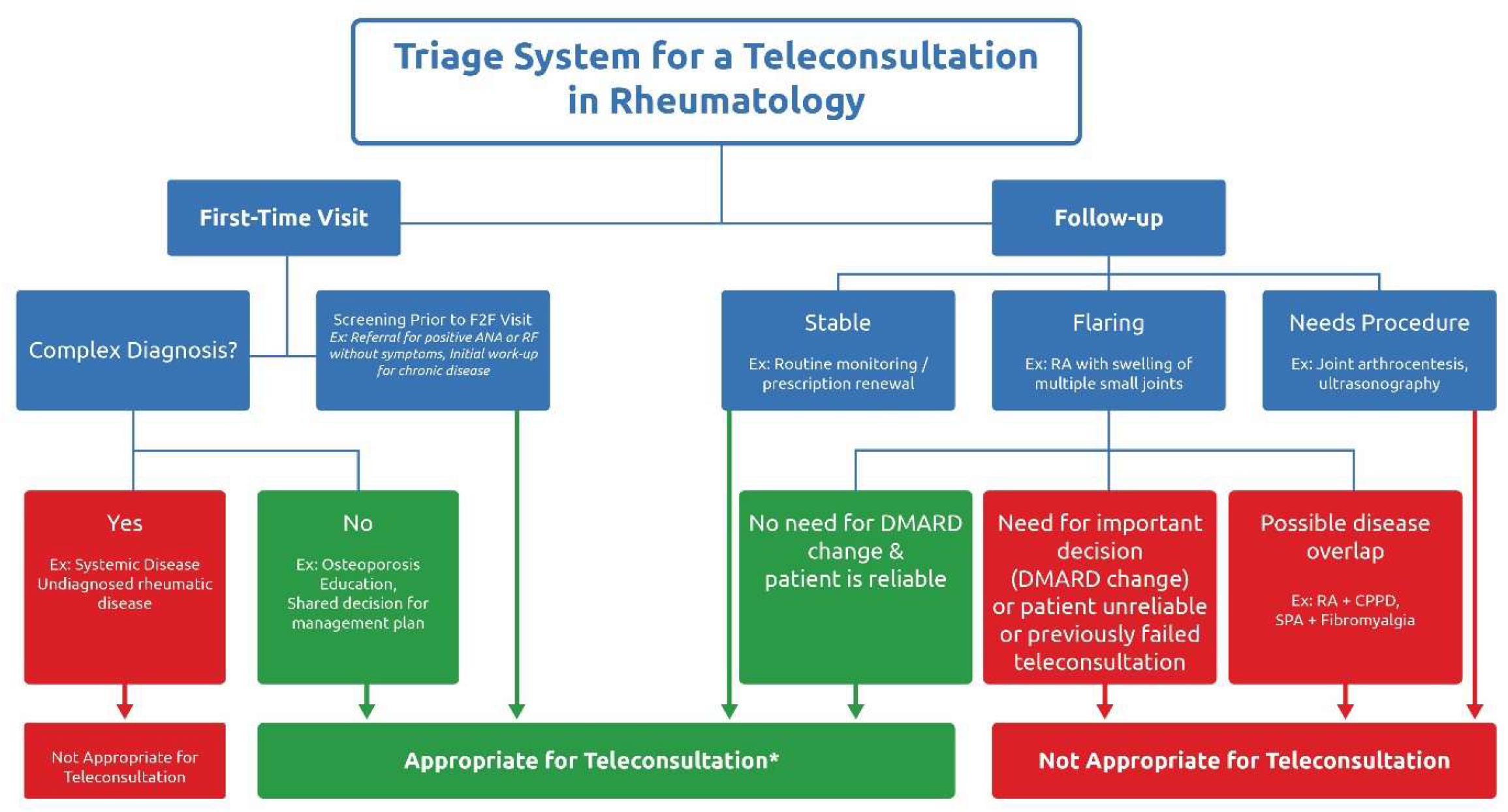
TRIAGE SYSTEM FOR A TELECONSULTATION IN RHEUMATOLOGY. Footnote: *Provided that the patient accepts teleconsultation

### Best Practice Statements

#### BPS 1: Informed Consent (LoE 5; LoA 7.90; Consensus 90.91%)

*Before the teleconsultation visit, an informed consent should be obtained from the patient, in writing or verbally; it should include an explanation, in a simple language, of the benefits and risks of telehealth encounters, as well as the conditions under which telehealth services may be terminated and a referral made to in-person care*.

Some telehealth providers may argue that when a patient electively chooses to visit a physician, whether it is a face-to-face visit or a teleconsultation, there is a “hidden” consent and that informed consent would only be required where the patient needs to undergo a procedure, or where patients are included in research trials. However, it should be emphasized that teleconsultation is a novel way of engaging with patients, and patients should be appropriately informed about the limitations and drawbacks of telehealth (e.g. misdiagnosis). Therefore, the focus is on “informed” consent, and patient consent should be obtained at least before the patient’s first teleconsultation. The explanation provided to the patient should include the nature of the telehealth encounter — including any technical limitations or potential for disruption and contingency plans, the protection of patient identifiable information, and billing information, if appropriate. It should also include the possibility to ask the patient to shift to an in-person visit if the physician considers that it would be more appropriate. At any point, before, during, or after the teleconsultation, the patient may withdraw consent from telehealth services.[14,23] While the standards and requirements of informed patient consent will likely vary depending on the jurisdiction, it is recommended to keep electronic records for informed consent.

#### BPS 2: Confidentiality (LoE 5; LoA 8.82; Consensus 100%)

*The use of telehealth services must ensure the patient’s information security and confidentiality*.

This should include proper and mandatory measures to ensure online information security (e.g., anti-hacking measures, password-protected access) and ensuring visual and auditory privacy in both the patient’s and the provider’s environment. Unless both parties explicitly agree upon it, the teleconsultation will not be recorded, whether by the patient or by the physician.

#### BPS 3: Documentation (LoE 5; LoA 8.77; Consensus 100%)

*The provision of telehealth services should be well documented in the patient’s file, similarly to any in-person medical visit. The decision to assess the patient remotely should be justified and recorded in the patient’s file*.

It is recommended that providers of telehealth services have access to the patient’s medical records, especially when the patient is not known to them. Furthermore, the reasoning behind whether or not to assess the patient via teleconsultation should be clearly documented, and the patient’s records should be updated accordingly.[14,24]

#### BPS 4: Shared decision and choice (LoE 5; LoA 8.36; Consensus 100%)

*The choice of using telehealth services should be based on a shared decision between the patient and the physician. Patients should have a choice of their provider of medical teleconsultation*.

The patients should be allowed to choose whether or not they want to engage with their rheumatologist via teleconsultation and to choose their telehealth provider without payers mandating the use of specified telehealth platforms or preferred providers with restrictive policies.[14]

#### BPS 5: Patient’s physical examination (LoE 2; LoA 7.77; Consensus 90.91%)

*Some parts of the physical exam might be performed remotely, like inspection and evaluation of the range of motion. The patient should be instructed on how to be prepared for a remote physical exam using appropriate educational material*.

While physical attendance of the patient is preferable in some cases — like a patient’s first consultation where an accurate examination and diagnosis is essential, some parts of the physical exam might be performed via teleconsultation. Physical examination is one of the pillars of medical reasoning in rheumatology; the inability to examine a patient might be viewed as a major obstacle. However, remote physical musculoskeletal examinations can be facilitated by the appropriate instructions to the patient with regards to what clothing to wear, the position of the camera, and the proper use of furniture (e.g., the use of a sofa or a chair to demonstrate the hip or knee range of motion). In some instances, assistance from a proxy person (e.g., a family member) can be requested for the physical exam. Physicians can also send a brochure or video tutorial regarding the maneuvers that will be performed for the physical examination to the patient before the teleconsultation.[25–29] It is recommended to translate and validate such tutorials in Arabic, with adaptation to local dialects where appropriate.[25,30]

#### BPS 6: Patient-reported outcomes (LoE 3; LoA 7.73; Consensus 100%)

*In some chronic rheumatic diseases, the use of patient-reported outcomes by means of self-completed questionnaires adapted for telehealth can help the physician make informed clinical decisions and improve the quality of care*.

In patients with an established disease, such as RA and spondyloarthritis, disease activity and functional status measures can be assessed before the teleconsultation using self-completed questionnaires of Patient-Reported Outcomes (PROs).[31,32] Such PROs can be transmitted with the rest of the patient’s medical file (laboratory results, radiology reports) before the teleconsultation to guide the physician on clinical decisions. For instance, studies have shown that among RA patients with low disease activity (LDA) or remission, a PROs-based telehealth follow-up for tight control of disease activity in RA can achieve similar disease control as a conventional outpatient follow-up.[33] RA Impact of Disease (RAID) score, in particular, has been shown to function well as a PRO in routine care, where patients with RAID <2 have a high likelihood of being in remission/ LDA (as per the disease activity score) and, if pre-screened, could avoid a clinic visit.[34] However, the validity of PROs measures needs to be evaluated in dedicated studies. In addition, several PROs measures should be translated and validated in Arabic and adapted to the cultural level of the patient when possible.

#### BPS 7: Safe Prescription (LoE 5; LoA 8.05; Consensus 95.45%)

*The prescription should be transmitted in a safe and confidential manner to the patient with a particular attention to avoiding abuse (of opioids and narcotics prescriptions in particular)*.

The risk of prescription abuse, particularly of opioids and narcotics, may be increased in a telehealth setting.[35,36] Therefore, an efficient tracking system of the prescriptions should be applied in the electronic health records and within the country’s legal framework where the rheumatologist is based. One such option is to have an electronic medical record connected with a platform that facilitates the timely teleconsultation between physician and patient while allowing the physician to write the prescription electronically. However, some countries do not allow electronic signatures for prescriptions, especially for opioids and narcotics, which needs to be considered.

#### BPS 8: Fees and reimbursement (LoE 5; LoA 8.45; Consensus 100%)

*The teleconsultation is subject to medical fees and reimbursement similarly to the in-person visit. Any fees should be set before the teleconsultation*.

The lack of a consensual reimbursement policy may be a significant deterrent to the adoption of telehealth. Therefore the physician and the legislator in the physician’s base country should agree on a fair and transparent fee for a teleconsultation; these fees should be communicated to the patient before the visit.[14]

Physicians should be aware of the fact that some patients might be unwilling to pay for an audio-only consultation, while some payers might be inclined to pay only a percentage of the conventional fee for teleconsultations that do not utilize video, e.g., 33% in UAE, 50% in Jordan, 80% in Bahrain (data provided by the TF during the Delphi rounds). Aside from the patient/payer perspective, some physicians might argue that teleconsultation can be more time-consuming than an in-person visit and that reimbursement should reflect the time spent with the patient. From a purely economic perspective, telehealth may decrease the indirect cost for the patient (e.g., by saving on travel costs, time taken off from work)[22] and for the physician (by reducing the number of no-shows and appointment cancellations).[37,38] By helping to maintain patients in remission or LDA through telemonitoring, the cost of managing chronic diseases would also be substantially reduced.[39]

#### BPS 9: Ethical considerations (LoE 5; LoA 8.59; Consensus 100%)

*Telehealth practice should conform to the same professional ethics that govern in-person care and comply with local jurisdictional laws and regulations of the physician’s location*.

Advancements in technology have facilitated the use of telehealth and Information Technology in the treatment or rehabilitation of diseases. However, increased use of technology is accompanied by threats to patients’ personal information. Therefore, special consideration to the ethical issues involved in telehealth practice, including technology, confidentiality and security, doctor-patient relationship, and informed consent, are crucial to guarantee safe use while maintaining the quality of healthcare services.[40]

#### BPS 10: Training of rheumatologists (LoE 5; LoA 8.18; Consensus 95.45%)

*The rheumatologists are encouraged to receive proper training through seminars, workshops, and conferences to familiarize with the advantages and disadvantages of telehealth and to acquire strategies about the most productive approach to remote medical care*.

Al-Samarraie *et al*. suggest that a lack of adequate training and familiarity with telehealth can be a barrier to the successful adoption. Therefore, it is recommended that rheumatologists and their staff educate themselves on all aspects of telehealth through continued professional development.[41] Although professional development and training can always add value to the physician, further evaluation of the effectiveness of training on the outcome of teleconsultations is necessary.

#### BPS 11: Improve infrastructure and promote equity (LoE 5; LoA 8.41; Consensus 100%)

*The technical infrastructure should be improved for patients and physicians to enable an efficient and equitable access to telehealth services across the countries and in vulnerable populations*.

Studies in the Middle East have shown that poor infrastructure was associated with low adoption of telehealth in the region.[42] This is especially true for patients living below the poverty line, as research indicates that access to telehealth seems to be a challenge for these vulnerable populations.[1] On the other hand, telehealth can also decrease inequities by offering a better chance of access and continuity of care for patients living in rural or remote areas. Rheumatologists should therefore identify limitations of the technical infrastructure in their clinic or region and take appropriate steps to attempt to overcome such limitations, when feasible. This will ensure that patients receive the same standard of care via a teleconsultation as they would during an in-clinic visit.[17] Having a secure platform that would allow the patient to make an appointment with their physician of choice, engage in teleconsultation, obtain the required prescription and diagnostic tests and pay the fees to the physician should be considered when possible. This could also include a rating system to rate the quality of the visit to help physicians improve the platform or mode of the consultation.

#### BPS 12: Support research projects in telehealth in the Arab Region (LoE 5; LoA 8.32; Consensus 95.45%)

*Local and regional research projects to assess the implementation of telehealth and the resulting disease outcomes in the Arab region are strongly encouraged*.

Recommendations from the Middle East also highlight the need for health initiatives to focus on health education and promotion to raise awareness of the benefits of telehealth services in the region.[41]

### Top Barriers And Facilitators To Telehealth

Voting on the top physician and patient-related barriers and facilitators to Telehealth in Rheumatology in the Arab region[1,6,44–48,20,22,25,33,37,41–43] revealed that concern about the quality of care and proper communication, internal and external technical difficulties were regarded as the top patient and physician-related barriers. Lack of alternatives when social distancing is required, and increased access to care were voted as the top physician and patient-related facilitators to telehealth, respectively (**Table 2)**.

**Table 2.**
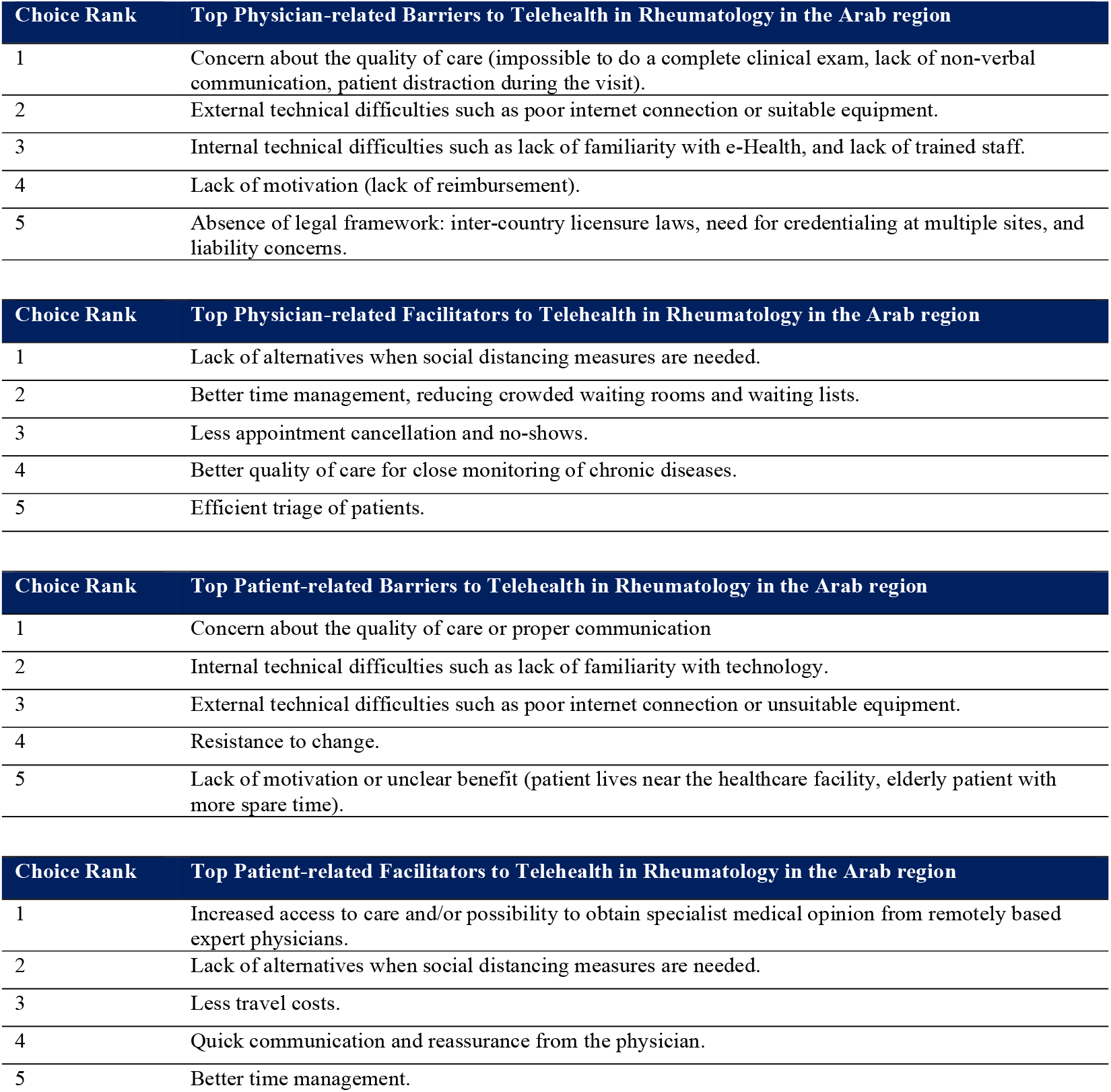
Top Physician- and Patient-related Barriers- and Facilitators to Telehealth in Rheumatology in the Arab region.

| Choice Rank | Top Physician-related Barriers to Telehealth in Rheumatology in the Arab region |
| --- | --- |
| 1 | Concern about the quality of care (impossible to do a complete clinical exam, lack of non-verbal communication, patient distraction during the visit). |
| 2 | External technical difficulties such as poor internet connection or suitable equipment. |
| 3 | Internal technical difficulties such as lack of familiarity with e-Health, and lack of trained staff. |
| 4 | Lack of motivation (lack of reimbursement). |
| 5 | Absence of legal framework: inter-country licensure laws, need for credentialing at multiple sites, and liability concerns. |

| Choice Rank | Top Physician-related Facilitators to Telehealth in Rheumatology in the Arab region |
| --- | --- |
| 1 | Lack of alternatives when social distancing measures are needed. |
| 2 | Better time management, reducing crowded waiting rooms and waiting lists. |
| 3 | Less appointment cancellation and no-shows. |
| 4 | Better quality of care for close monitoring of chronic diseases. |
| 5 | Efficient triage of patients. |

| Choice Rank | Top Patient-related Barriers to Telehealth in Rheumatology in the Arab region |
| --- | --- |
| 1 | Concern about the quality of care or proper communication |
| 2 | Internal technical difficulties such as lack of familiarity with technology. |
| 3 | External technical difficulties such as poor internet connection or unsuitable equipment. |
| 4 | Resistance to change. |
| 5 | Lack of motivation or unclear benefit (patient lives near the healthcare facility, elderly patient with more spare time). |

| Choice Rank | Top Patient-related Facilitators to Telehealth in Rheumatology in the Arab region |
| --- | --- |
| 1 | Increased access to care and/or possibility to obtain specialist medical opinion from remotely based expert physicians. |
| 2 | Lack of alternatives when social distancing measures are needed. |
| 3 | Less travel costs. |
| 4 | Quick communication and reassurance from the physician. |
| 5 | Better time management. |

### Triage System For Teleconsultation In Rheumatology

A Triage System for a Teleconsultation in Rheumatology (**Figure 3**) was developed to assist rheumatologists in assessing whether telehealth is suitable in each situation, based on the complexity of the diagnosis, the patient’s clinical status, and disease prognosis. Rheumatologists need to use their professional experience and judgment to assess whether telehealth is suitable in each situation while ensuring that the services provided comply with the laws and regulations of their respective countries. In addition, a practical toolkit for the possible implementation of teleconsultation in Rheumatology was developed to provide guidance on how to translate theory into practice, highlighting the activities to perform before, during, and after the teleconsultation (**Figure 4**).

**FIGURE 4.**
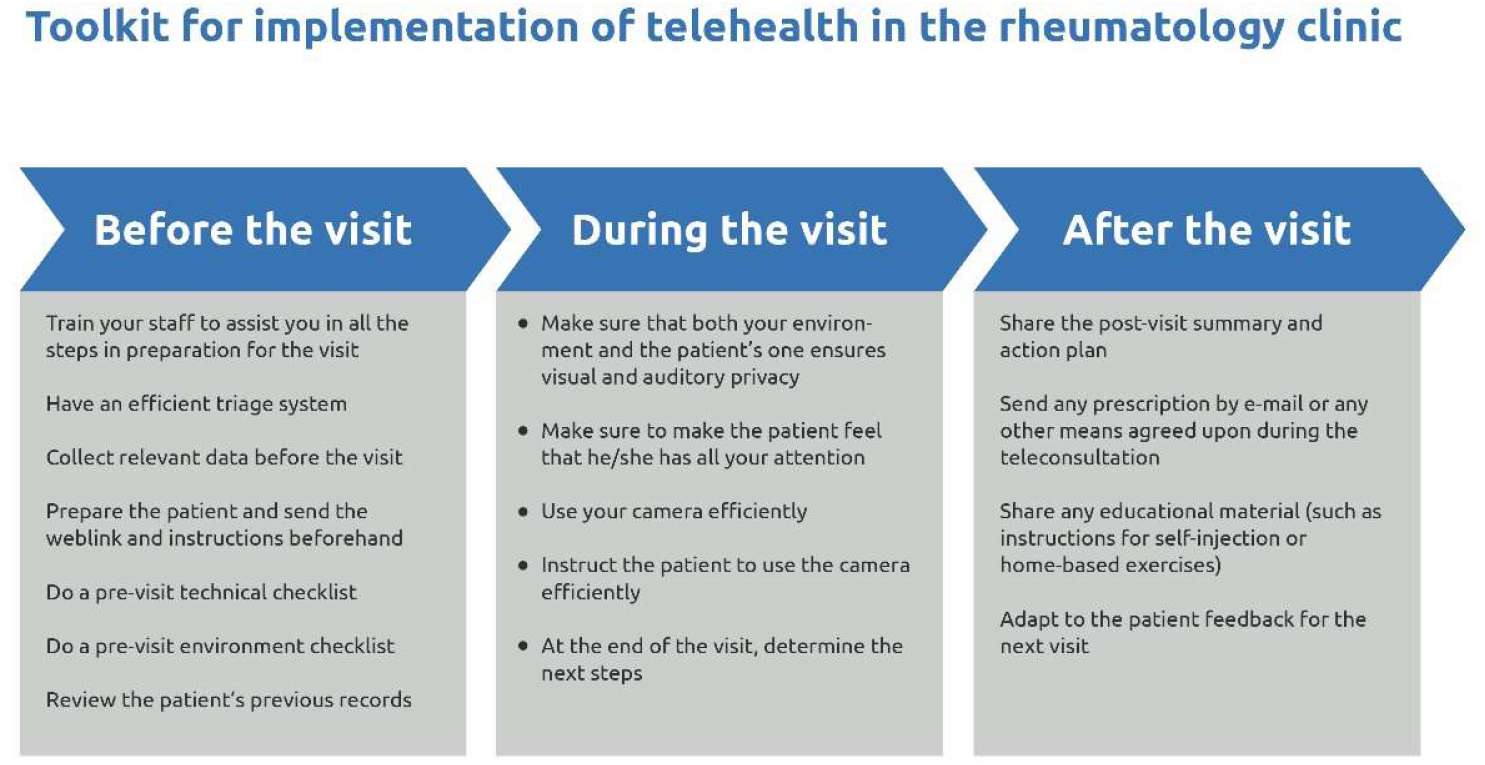
TOOLKIT FOR THE IMPLEMENTATION OF TELEHEALTH IN THE RHEUMATOLOGY CLINIC. **\*Technical checklist**: Check if your device and browser support the telehealth platform Check if your internet is stable Check sound and webcam on your device Have a backup plan if the connection fails Take time to familiarize yourself with the telehealth platform and its functions Ensure all your devices are charged and have the chargers handy in case your battery drain; Close other apps/programs before the visit to improve speed/connectivity \*\***Environment checklist**: Set up your workspace so that you are front-lit, the camera view covers approximately your head and shoulders, and the camera is at eye height Make sure the background is not distracting You might consider using a virtual background with your hospital logo or design Dress professionally

## DISCUSSION

The ArLAR BPG for telehealth were developed to inform the rheumatologists about the advantages and limitations of telehealth and to provide them with a series of strategies to practice telehealth in the rheumatology clinic.

The need for practical ways to provide remote health care has been highlighted during the COVID-19 pandemic, where social distancing and potential risks to patient safety prohibit regular in-person consultations. In that setting, telehealth can be a valuable tool for managing patients with rheumatic disease, especially more vulnerable patients with co-morbidities and other risk factors.[49] In addition, recent studies suggest that telehealth services could positively affect factors such as disease activity, medication adherence, physical activity, and self-efficacy levels in patients with RA, provided these interventions are well-designed, versatile, and adaptive.[50]

While some studies have found telehealth to be generally effective for the diagnosis and self-management of rheumatic disease, with mostly positive patient and provider satisfaction,[15,50] others have found that the use of telerheumatology during the COVID-19 pandemic was associated with diagnostic delay, reduced likelihood of changing existing immunosuppressive therapy, earlier requirement for review and a lower likelihood of discharge, even though it led to improved appointment attendance.[51]

This highlights the notion that the appropriateness of telerheumatology in patients’ care may vary widely, differing by age, phase of illness, severity of symptoms, and rheumatic disease type. Furthermore, the effects of telerheumatology on patient access and the outcome of rheumatic disease across race, ethnicity, or socioeconomic status have not been explicitly studied in literature and may be difficult to predict.[52] In a study evaluating the patient’s feelings about changing from a face-to-face consultation to a virtual one, only 76% of patients had the means to access a teleconsultation. The proportion of internet access and the agreement for a teleconsultation decreased in patients over 70 years old. The factors associated with an acceptance for teleconsultation were the significant distance from the consultation site and the higher level of education.

The TIROL Steering Committee acknowledges the significant disparity between countries in terms of available laws and legislation that manages telehealth and that any guidelines will always be subject to compliance with the applicable laws and regulations of each ArLAR country. Rheumatologists are encouraged to actively participate in the effort to sensitize legislators to the interest and benefit of telerheumatology. Focusing on the patient- and physician-related barriers in the Arab region can assist authorities in establishing appropriate strategies for promoting telehealth. Moreover, the committee acknowledges the disparity in internet accessibility, electricity stability, and technology skills across the Arab countries, which are critical to consider when deciding whether telehealth is an appropriate method of providing health care. In the absence of clear evidence, rheumatologists should use their professional experience and judgment to assess whether telehealth is suitable in each situation while always prioritizing security and confidentiality considerations.

In summary, the ArLAR BPG provide a consensual framework for the application of telehealth in rheumatology practice. Most of the statements were based on a low level of evidence and expert opinion, highlighting the need for further dedicated research to recognize the advantages and limitations of telehealth in rheumatology and for a subsequent update of these BPG in the future. The ArLAR BPG for telehealth are not intended to compel the use of telehealth in the same manner for every patient in every case; but are rather designed to provide the rheumatologists with a series of strategies about the most reliable, productive, and rational approaches to apply telehealth in the rheumatology setting, and principally in context of pressin epidemic restrictions.

## Data Availability

Extra data regarding the details about all three Delphi rounds are available in the supplementary material.

## Authors statement

Nelly Ziade, Ihsane Hmamouchi, Lina el Kibbi, Melissa Daou, Nizar Abdulateef and Fatemah Abutiban constituted the steering committee which designed the study, wrote the protocol and supervised the study.

Melissa Daou and Nelly Ziade performed the literature review.

Nelly Ziade and Ilanca Fraser analyzed the results of the Delphi rounds wrote the first manuscript draft. All the authors made substantial contributions to work and participated in the three Delphi rounds.

All the authors participated to the draft of the manuscript and revised the final submitted document for intellectual content.

All the authors approved the version to be published and agreed to be accountable for all aspects of the work.

## Acknowledgments

The authors would like to acknowledge the following people for their contribution to this study: Dr Christine Peoples, the patients, regulators, payers and rheumatologists who participated in the Delphi survey, including Dr Nazih El Kouartey and Dr Lara Haddadin. We also thank Pfizer Gulf FZ LLC for providing an unrestricted educational grant to fund the medical writing and Delphi platform for this project and Taylor Wessing© for providing legal advice.

