## Supplementary Table 1 for "Telehealth in Rheumatology: The 2021 Arab League of Rheumatology Best Practice Guidelines"

**Table 1.** General Principles and Best Practice Guidelines statements for Telehealth in Rheumatology

|  | |  | Round 1 | | Round 2 | | Round 3 | |
| --- | --- | --- | --- | --- | --- | --- | --- | --- |
| General Principles | | LoE* | Consensus**  (%) | LoA***  Arithmetic Mean (SD)/ Median | Consensus**  (%) | LoA ***  Arithmetic Mean (SD)/ Median | Consensus**  (%) | LoA***  Arithmetic Mean (SD)/ Median |
| A  Definition of a teleconsultation | A rheumatology teleconsultation is a synchronous exchange of medical information‡ between a patient and a rheumatologist via audio or audio-visual electronic communication, to improve the patient’s health status. | 5 | 86.37 | 7.64 (0.98)  7 | 100 | 8.14 (0.77)  8 | 100 | 8.14 (0.77)  8 |
| B  Access and Continuity of Care | Telehealth may improve the access and continuity of care for patients with rheumatic diseases who are home-bound, live in remote areas or under-served communities, or who need to adhere to social distancing restrictions. | 3 | 100 | 8.41(0.80)  9 | 100 | 8.41 (0.80)  9 | 100 | 8.41 (0.80)  9 |
| C  Improving disease outcomes | Telehealth can help some patients adhere to the management plan and this is likely to improve disease outcomes in some selected disease states. | 2 | 81.82 | 7.18 (1.26)  7 | 90,91 | 7.77 (1.04)  8 | 86.36 | 7.86 (1.21)  8 |
| D  Quality of medical care | Rheumatologists need to use professional experience and judgment to assess whether telehealth is suitable in each situation. | 5 | 95,45 | 8.14 (0.89)  8 | 95,45 | 8.14 (0.89)  8 | 95.45 | 8.14 (O.89)  8 |
| Best Practice Statements | | | | | | | | |
| 1  Informed consent | Before the teleconsultation visit, an informed consent should be obtained from the patient, in writing or verbally; it should include an explanation, in a simple language, of the benefits and risks of telehealth encounters, as well as the conditions under which telehealth services may be terminated and a referral made to in-person care. | 5 | 72.73 | 7.00 (1.51)  8 | 90,91 | 7.86 (1.28)  8 | 90.91 | 7.86 (1.28)  8 |
| 2  Confidentiality | The use of telehealth services must ensure the patient’s information security and confidentiality. | 5 | 100 | 8.73 (0.54)  9 | 100 | 8.82 (0.50)  9 | 100 | 8.82 (0.50)  9 |
| 3  Documentation | The provision of telehealth services should be well documented in the patient’s file, similarly to any in-person medical visit. The decision to assess the patient remotely should be justified and recorded in the patient’s file. | 5 | 100 | 8.77 (0.43)  9 | 100 | 8.77 (0.43)  9 | 100 | 8.77 (0.43)  9 |
| 4  Shared decision and choice | The choice of using telehealth services should be based on a shared decision between the patient and the physician. Patients should have a choice of their provider of medical teleconsultation. | 5 | 100 | 8.05 (0.83)  8 | 100 | 8.36 (0.73)  8.5 | 100 | 8.36 (0.73)  8.5 |
| 5  Patient’s physical examination | Some parts of the physical exam might be performed remotely, like inspection and evaluation of the range of motion. The patient should be instructed on how to be prepared for a remote physical exam, using appropriate educational material. | 2 | 77.27 | 7.27 (1.29)  7 | 90,91 | 7.77 (1.02)  8 | 90,91 | 7.77 (1.02)  8 |
| 6  Patient-reported outcomes | In some chronic rheumatic diseases, the use of patient-reported outcomes by means of self-completed questionnaires adapted for telehealth can help the physician make informed clinical decisions and improve the quality of care. | 3 | 81.82 | 7.32 (1.10)  7 | 100 | 7.73 (0.77)  8 | 100 | 7.73 (0.77)  8 |
| 7  Safe Prescription | The prescription should be transmitted in a safe and confidential manner to the patient with a particular attention to avoiding abuse (of opioids and narcotics’ prescriptions in particular). | 5 | 95,45 | 8.05 (0.90)  8 | 95,45 | 8.05 (0.90)  8 | 95.45 | 8.05 (0.90)  8 |
| 8  Fees and reimbursement | The teleconsultation is subject to medical fees and reimbursement similarly to the in-person visit. Any fees should be set before the teleconsultation. | 5 | 77.27 | 7.45 (1.64)  8 | 100 | 8.23 (0.73)  8 | 100 | 8.45 (0.60)  8.5 |
| 9  Ethical considerations | Telehealth practice should conform to the same professional ethics that govern in-person care and comply with local jurisdictional laws and regulations of the physician’s location. | 5 | 100 | 8.59 (0.60)  9 | 100 | 8.59 (0.60)  9 | 100 | 8.59 (0.60)  9 |
| 10  Rheumatologist training | The rheumatologists are encouraged to receive proper training through seminars, workshops, and conferences to familiarize with the advantages and disadvantages of telehealth and to acquire strategies about the most productive approach to remote medical care. | 5 | 95,45 | 8.18 (1.05)  8.5 | 95,45 | 8.18 (1.05)  8.5 | 95.45 | 8.18 (1.05)  8.5 |
| 11  Technical infrastructure and equity | The technical infrastructure should be improved for patients and physicians to enable an efficient and equitable access to telehealth services across the countries and in vulnerable populations. | 5 | 100 | 8.41 (0.73)  9 | 100 | 8.41 (0.73)  9 | 100 | 8.41 (0.73)  9 |
| 12  Research | Local and regional research projects to assess the implementation of telehealth and the resulting disease outcomes in the Arab region are strongly encouraged. | 5 | 95,45 | 8.32 (1.04)  9 | 95,45 | 8.32 (1.04)  9 | 95.45 | 8.32 (1.04)  9 |

** LoE : Level of Evidence according to* the Oxford Centre for Evidence-Based Medicine

***Consensus : % of votes with a score from 7 to 9*

**** LoA : Level of agreement from 1 to 9, with 9 being the highest agreement.*

‡Exchange of medical information includes medical history, physical exam, review of paraclinical results and final prescription.
